# A Comparison of Changes in Venous Lactate and Haematocrit during Fluid Resuscitation of Dengue Haemorrhagic Fever

**DOI:** 10.1101/2022.11.14.22282277

**Authors:** W D Dilshan Priyankara, D G N Samarutilake, Sameera Viswakula, E M Manoj, Ananda Wijewickrama, Nilanka Perera, J K P Wanigasuriya

## Abstract

**Background:** Dengue haemorrhagic fever (DHF) causes significant morbidity and mortality. Judicious fluid resuscitation is the cornerstone of managing vascular leakage in DHF and haematocrit (HCT) measurement is used to monitor the response to fluid resuscitation. Serum lactate level is a measure of tissue perfusion which can be a useful parameter to monitor adequate fluid therapy. The usefulness of lactate in the management of DHF is poorly investigated.

**Methodology and Principal Findings:** A prospective observational study was conducted in two treatment centers in Sri Lanka recruiting 162 DHF patients, to study the correlation of venous lactate and HCT during fluid resuscitation. Patients were recruited within 12 hours of diagnosis of the critical phase and venous lactate level was measured at each time of performing HCT, using a pre-validated handheld lactate analyzer. Median lactate level was 1.3 (range 0.3 - 6 mmol/L) in the study population and 154 (95.2%) patients had median lactate levels less than 2 mmol/L. The HCT values in the study participants ranged from 28 to 62, with a median value of 43. There was no statistically significant correlation between the lactate and HCT values obtained at the same time. In addition, a statistically significant reduction in venous lactate was not observed following administration of fluid boluses. However, HCT reduction expected by administration of the fluid boluses was seen following dextran and crystalloid/dextran combination. Capillary HCT increased following blood transfusion. The highest lactate level measured in a patient was associated with an increase in hospital stay.

**Conclusions:** This study concludes that venous lactate is not an appropriate parameter to monitor response to fluid therapy in uncomplicated DHF.

**Author Summary:** Dengue viral infection causes asymptomatic disease to severe haemorrhagic fever causing organ failure and death. Severe manifestations occur due to fluid extravasation during the critical phase of the illness and these patients with dengue haemorrhagic fever (DHF) require close monitoring and guided fluid therapy. Adequacy of fluid resuscitation is guided by capillary haematocrit (HCT) measurement. However, HCT does not reflect the tissue perfusion. Venous lactate is a reliable measure of tissue perfusion is circulatory collapse. Lactate is known to be a useful marker in identifying severe dengue disease. The usefulness of venous lactate to predict tissue perfusion during fluid resuscitation of DHF has not been performed. The present study was done to identify the usefulness of venous lactate measured by a hand-held lactate analyser in fluid resuscitation of DHF and to find the correlation of HCT and lactate values. Results revealed that uncomplicated DHF patients did not have significantly elevated lactate levels and the HCT and lactate levels performed at the same time did not correlate. Highest lactate level measured in a patient was associated with a longer hospital stay. Therefore, venous lactate is not an appropriate marker to guide fluid therapy in uncomplicated DHF.

## Introduction

Dengue is the most common mosquito-borne viral infection worldwide [1][2] and it is a major public health concern in developing countries such as Sri Lanka. Globally, the number of reported dengue cases have increased from 2.4 million in 2010 to 5.2 million in 2019 and there has been a sharp rise during the last few years [3]. Estimated disease burden is much higher and seventy percent of apparent infections occur in Asia [4]. In Sri Lanka, dengue infection has spread throughout the country with all four serotypes circulating during the last 30 years [5]. According to the World Health Organization (WHO), around 20000 patients die from dengue infection each year globally [1].

Dengue fever (DF) is a self-limiting febrile illness in the majority, but some patients can progress into a severe and potentially life-threatening illness; dengue hemorrhagic fever (DHF) and dengue shock syndrome (DSS). Fluid leakage due to increased vascular permeability is the main pathophysiological feature in DHF or DSS [6]. The mechanism of increased vascular permeability and fluid leakage in dengue is not well established. The degree of fluid leakage determines the severity of DHF and the hypovolaemia results in poor circulation leading to organ dysfunction and multi-organ failure [7]. Management of DHF and DSS has been challenging and the available guidelines are mainly based on expert opinion and consensus statements [8].

Management of DHF and DSS is guided by clinical parameters (pulse, blood pressure and urine output) and the measured capillary haematocrit (HCT). The haemoconcentration occurring as the result of vascular leakage causes a rise in the HCT which is used to guide fluid resuscitation. However, it is questionable whether the end organ perfusion is reflected accurately by the HCT measurement alone. Serum lactate is a well-known marker of tissue perfusion [9]. Therefore, serum lactate could potentially be a useful indicator to identify effective tissue perfusion in the management of DHF and DSS. In addition, use of capillary lactate available at bedside, could be an additional readily available parameter during resuscitation of DHF or DSS in the wards. Previous studies have demonstrated that serum lactate can be used to identify severe dengue infection either alone or in combination with additional parameters[10–14] However, the usefulness of lactate to guide fluid therapy has not been identified. Identifying the venous lactate trends in DHF and DSS and analyzing the correlation with HCT is extremely useful. The present study was designed to compare venous lactate values with venous HCT in DHF and DSS and identify the usefulness of venous lactate as a parameter during fluid resuscitation.

## Materials and Methods

### Study design and setting

A prospective observational study was conducted in the dengue treatment units of National Institute of Infectious Diseases, Angoda, Sri Lanka and Colombo South Teaching Hospital, Sri Lanka from 01.09.2019 to 30.01.2020. Study population included patients with confirmed dengue fever who developed DHF or DSS during the hospital stay. Dengue viral infection was confirmed by a positive dengue NS1 antigen or positive dengue serology (IgM). Patients less than 18 years of age, patients with expanded dengue syndrome without evidence of fluid leakage and patients with morbidities such as chronic kidney disease (stage 3 or above) or chronic liver disease were excluded. In addition, pregnant mothers and patients with co-infections were excluded from the study.

### Study procedure

All patients who developed DHF or DSS either on-admission or after admission to the study setting, were enrolled within 12 hours of diagnosis of the critical (leaking) phase. Written informed consent was obtained from the patients by the study investigators. All patients were managed according to the national guideline published by the Ministry of Health, Sri Lanka and the critical phase monitoring chart was maintained. The time of HCT measurement was decided by the treating medical team according to the guideline. Venous lactate level was performed each time of performing HCT using a pre-validated handheld lactate analyzer (Lactate Scout). Serial lactate measurements (3-4 hourly) were performed along with HCT until the end of the critical phase as described in the national guideline. Baseline characteristics of the patients, laboratory parameters, clinical parameters, amount of fluid required for resuscitation, requirement of colloids and blood products were obtained from the patient’s clinical notes. Complications such as bleeding, secondary sepsis, fluid overload, transaminitis (liver enzymes greater than 3 times upper limit of normal) and the outcome (ICU admission, death, length of ICU and hospital stay) were recorded. Serial lactate values and HCT values were recorded on the dengue monitoring chart.

### Statistical analysis

Data was entered into an MS Excel sheet and double checked for validity. Data analysis was performed using IBM® SPSS® Statistics version 22. Demographic characteristics and other parameters were analyzed using descriptive statistical methods and presented as percentages or mean/median as appropriate. Pearson’s correlation coefficient was performed to find the correlation between the individual peak values of liver enzymes and venous lactate values. Paired sample t-test to compare pre and post values of HCT and lactate.

### Ethical consideration

The study was approved by the Ethical review committee of the Faculty of Medical Sciences, University of Sri Jayewardenepura (ERC Protocol No: 78/17) and the permission was obtained from both institutions to carry out the study.

## Results

### Characteristics of the study population

A total of 162 patients with DHF fulfilling inclusion criteria were recruited to the study. Majority of participants (n=135, 83.3%) did not have any underlying chronic medical illnesses and they were not on long term medical treatment (Table 1). Fever was the commonest symptom on admission followed by headache, arthralgia and myalgia. Median day of onset of the critical phase was 4 (IQR 3-5). The mean platelet count was 48.8 ×10^3^/µL (SD 36.6). Majority of participants (n=124, 76.5%) had free fluid in the abdomen denote leaking and only 24 (14.8%) had a HCT rise above 20% (Table 1).

**Table 1:**
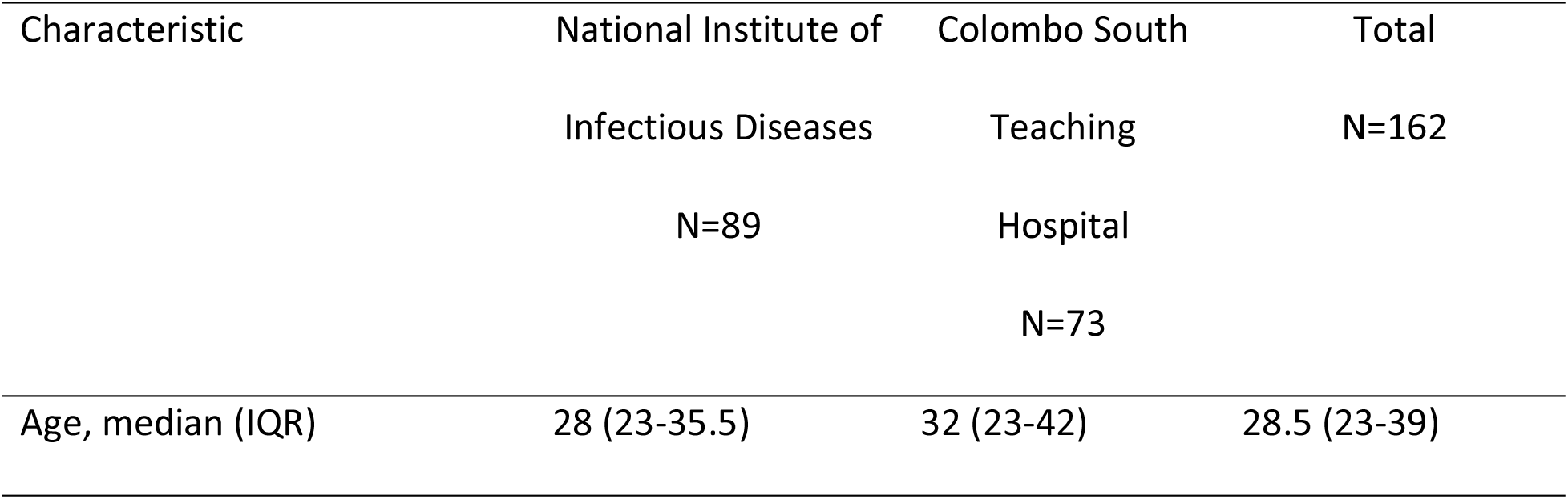
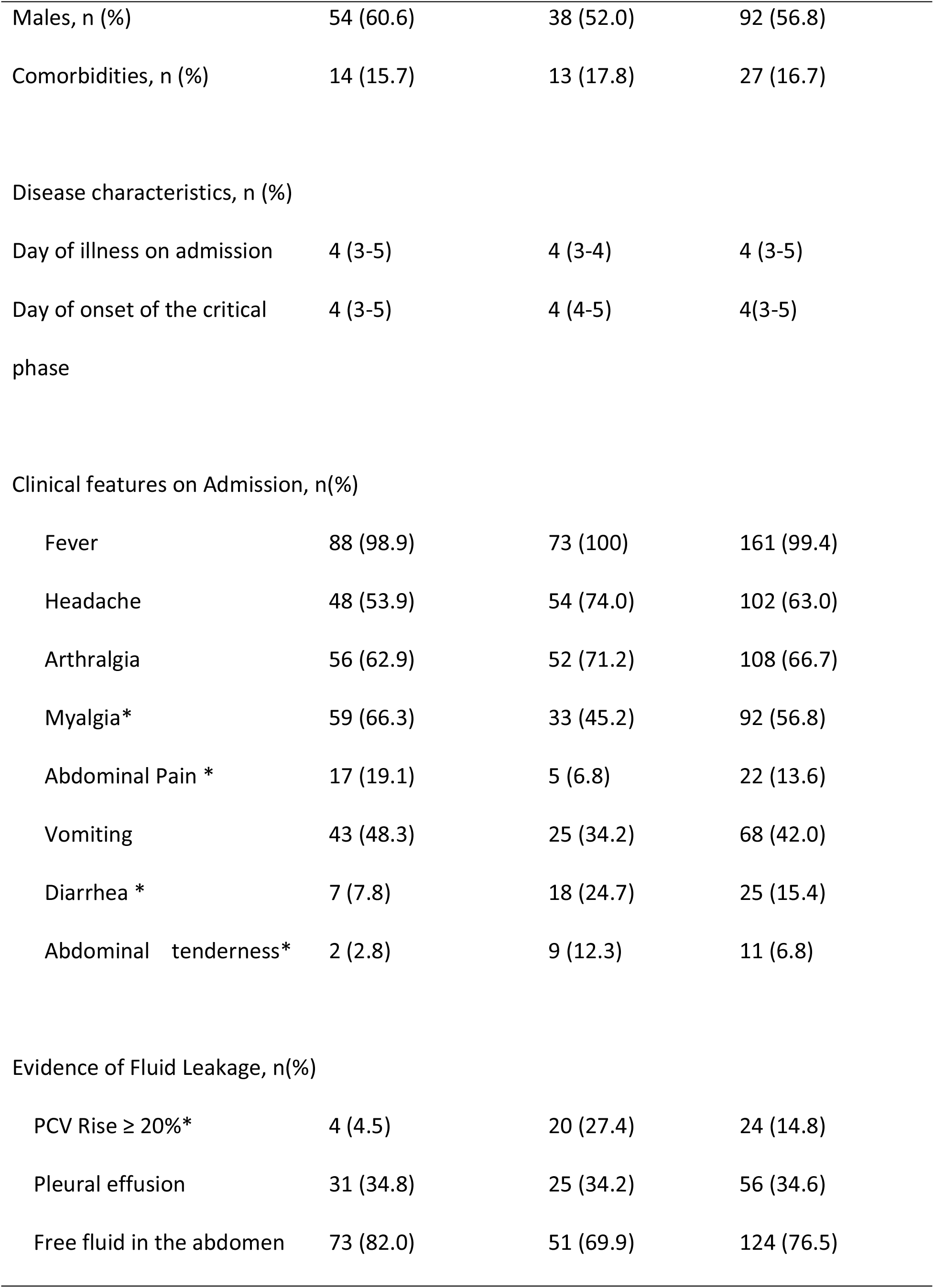
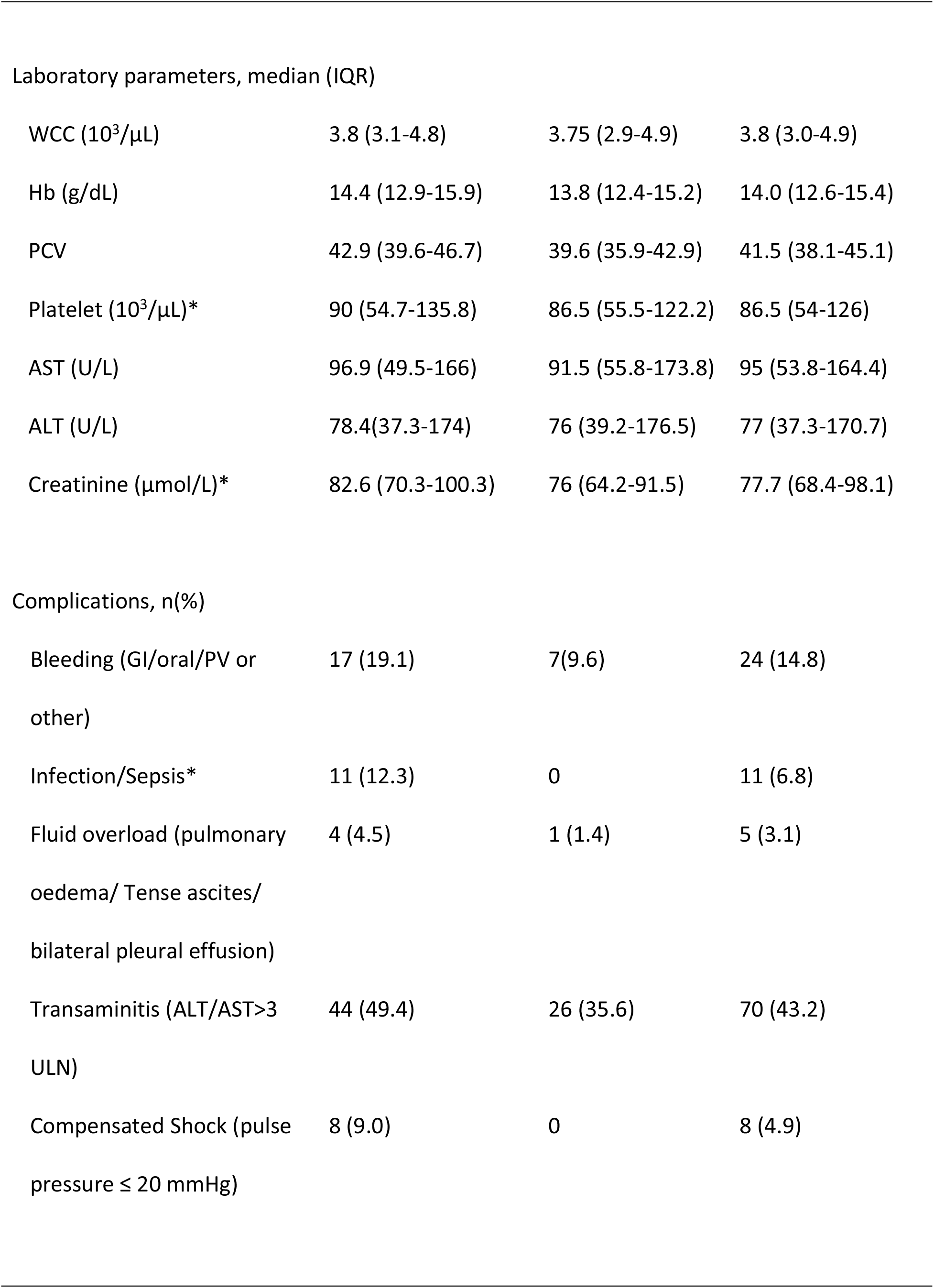
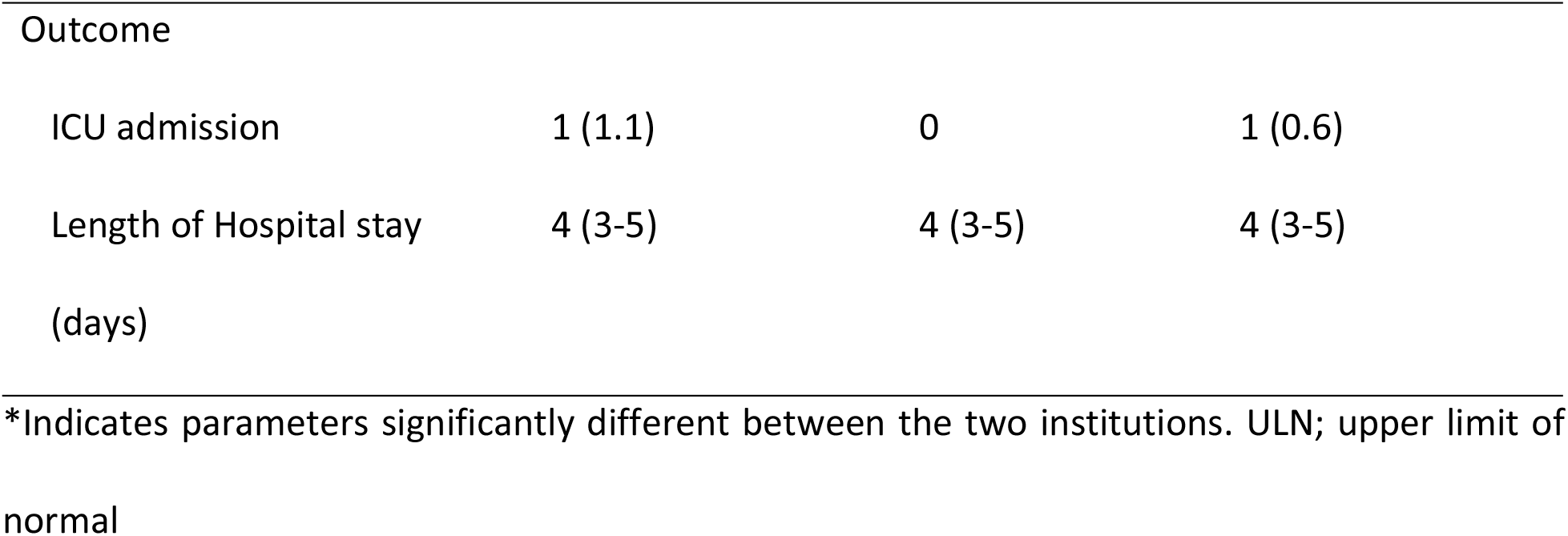
Clinical characteristics and the investigation findings of the study population.

| Characteristic | National Institute of<br>Infectious Diseases<br>N=89 | Colombo South<br>Teaching<br>Hospital<br>N=73 | Total<br>N=162 |
| --- | --- | --- | --- |
| Age, median (IQR) | 28 (23-35.5) | 32 (23-42) | 28.5 (23-39) |
| Males, n (%) | 54 (60.6) | 38 (52.0) | 92 (56.8) |
| Comorbidities, n (%) | 14 (15.7) | 13 (17.8) | 27 (16.7) |
| Disease characteristics, n (%) |  |  |  |
| Day of illness on admission | 4 (3-5) | 4 (3-4) | 4 (3-5) |
| Day of onset of the critical phase | 4 (3-5) | 4 (4-5) | 4(3-5) |
| Clinical features on Admission, n(%) |  |  |  |
| Fever | 88 (98.9) | 73 (100) | 161 (99.4) |
| Headache | 48 (53.9) | 54 (74.0) | 102 (63.0) |
| Arthralgia | 56 (62.9) | 52 (71.2) | 108 (66.7) |
| Myalgia* | 59 (66.3) | 33 (45.2) | 92 (56.8) |
| Abdominal Pain * | 17 (19.1) | 5 (6.8) | 22 (13.6) |
| Vomiting | 43 (48.3) | 25 (34.2) | 68 (42.0) |
| Diarrhea * | 7 (7.8) | 18 (24.7) | 25 (15.4) |
| Abdominal tenderness* | 2 (2.8) | 9 (12.3) | 11 (6.8) |
| Evidence of Fluid Leakage, n(%) |  |  |  |
| PCV Rise ≥ 20%* | 4 (4.5) | 20 (27.4) | 24 (14.8) |
| Pleural effusion | 31 (34.8) | 25 (34.2) | 56 (34.6) |
| Free fluid in the abdomen | 73 (82.0) | 51 (69.9) | 124 (76.5) |
| Laboratory parameters, median (IQR) |  |  |  |
| WCC (10<sup>3</sup>/μL) | 3.8 (3.1-4.8) | 3.75 (2.9-4.9) | 3.8 (3.0-4.9) |
| Hb (g/dL) | 14.4 (12.9-15.9) | 13.8 (12.4-15.2) | 14.0 (12.6-15.4) |
| PCV | 42.9 (39.6-46.7) | 39.6 (35.9-42.9) | 41.5 (38.1-45.1) |
| Platelet (10<sup>3</sup>/μL)* | 90 (54.7-135.8) | 86.5 (55.5-122.2) | 86.5 (54-126) |
| AST (U/L) | 96.9 (49.5-166) | 91.5 (55.8-173.8) | 95 (53.8-164.4) |
| ALT (U/L) | 78.4(37.3-174) | 76 (39.2-176.5) | 77 (37.3-170.7) |
| Creatinine (μmol/L)* | 82.6 (70.3-100.3) | 76 (64.2-91.5) | 77.7 (68.4-98.1) |
| Complications, n(%) |  |  |  |
| Bleeding (GI/oral/PV or<br>other) | 17 (19.1) | 7(9.6) | 24 (14.8) |
| Infection/Sepsis* | 11 (12.3) | 0 | 11 (6.8) |
| Fluid overload (pulmonary<br>oedema/ Tense ascites/<br>bilateral pleural effusion) | 4 (4.5) | 1 (1.4) | 5 (3.1) |
| Transaminitis (ALT/AST>3<br>ULN) | 44 (49.4) | 26 (35.6) | 70 (43.2) |
| Compensated Shock (pulse<br>pressure ≤ 20 mmHg) | 8 (9.0) | 0 | 8 (4.9) |

| Outcome |  |  |  |
| --- | --- | --- | --- |
| ICU admission | 1 (1.1) | 0 | 1 (0.6) |
| Length of Hospital stay<br>(days) | 4 (3-5) | 4 (3-5) | 4 (3-5) |

\*Indicates parameters significantly different between the two institutions. ULN; upper limit of normal

### Lactate levels in DHF

Venous lactate levels were measured serially in the DHF patients at each time point of testing HCT. There were 6 measurements of lactate on average for each patient. The highest lactate level recorded in a study participant was 6 mmol/L and the lowest value was 0.3 mmol/L with a median value of 1.3 mmol/L during the critical phase. The median lactate value for each patient was calculated and the distribution is given in Table 2. The HCT values in the study participants ranged from 28 to 62, with a median value of 43. Median HCT for each participant was calculated. There were 6 (3.7%) participants with a median HCT in the 25-35 range, 113 (69.7%) in the 36-45 range and 43 (26.5%) having a median HCT more than 45.

**Table 2:**
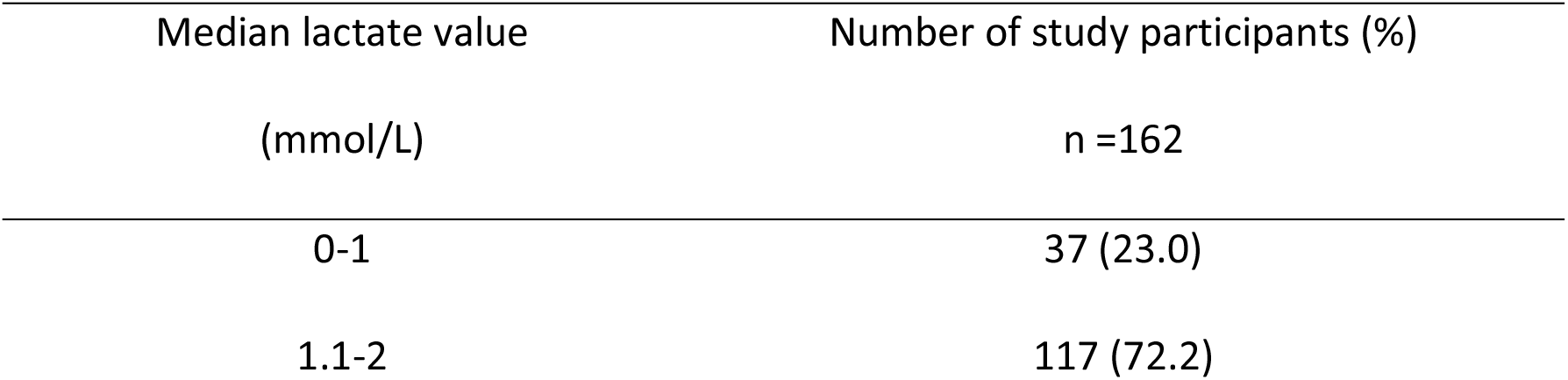
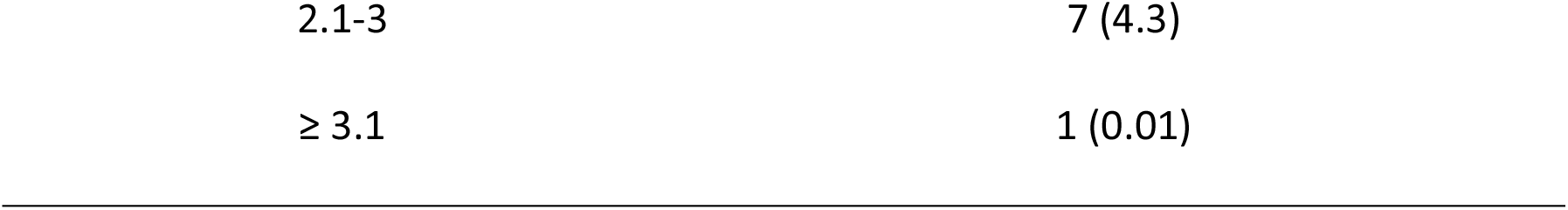
Distribution of venous lactate in the study participants during critical phase.

| Median lactate value<br>(mmol/L) | Number of study participants (%)<br>n =162 |
| --- | --- |
| 0-1 | 37 (23.0) |
| 1.1-2 | 117 (72.2) |
| 2.1-3 | 7 (4.3) |
| $\geq 3.1$ | 1 (0.01) |

There were 967 total lactate and corresponding HCT values for all participants analyzed in the study. There was no statistically significant correlation between the lactate and HCT values obtained at the same time (Figure 1).

**Figure 1:**
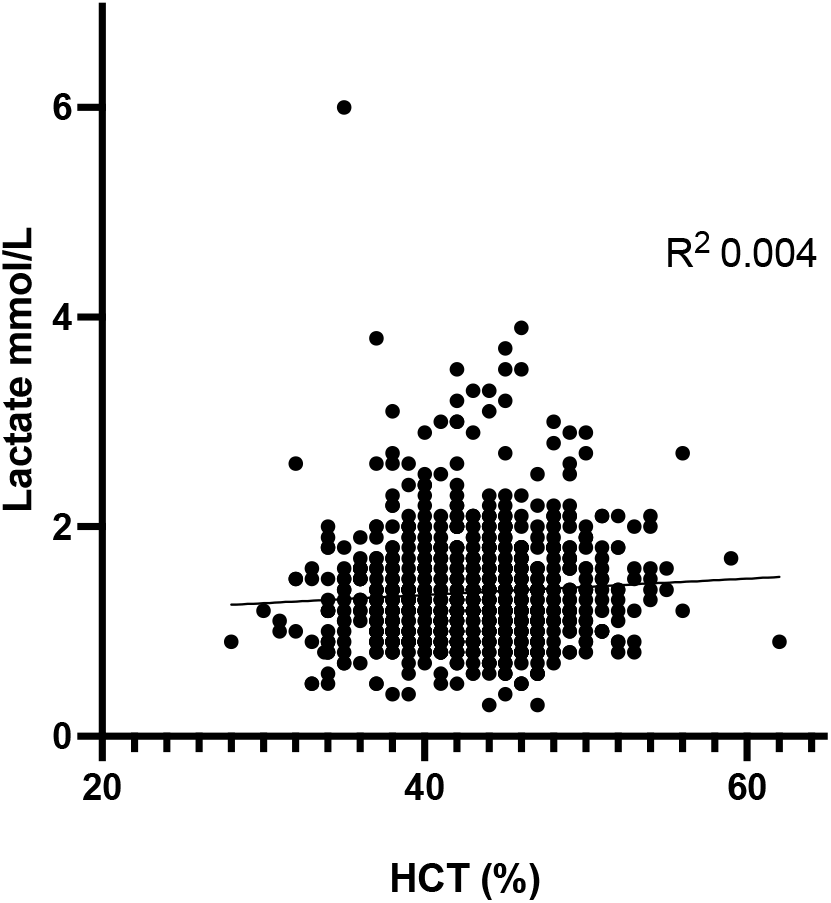
Association of lactate and HCT values. Lactate values (mmol/L) and corresponding HCT (%) values were plotted in a dot plot graph. Figure represents 967 corresponding values. Regression co-efficient of determination (R2) 0.004.

Pulse pressure reduction is a reflection of the worsening haemodynamic status in DHF. The highest lactate or the highest HCT recorded in a patient was not associated with the lowest pulse pressure detected. However, the highest lactate level measured in a patient was associated with an increase in hospital stay (Figure 2).

**Figure 2:**
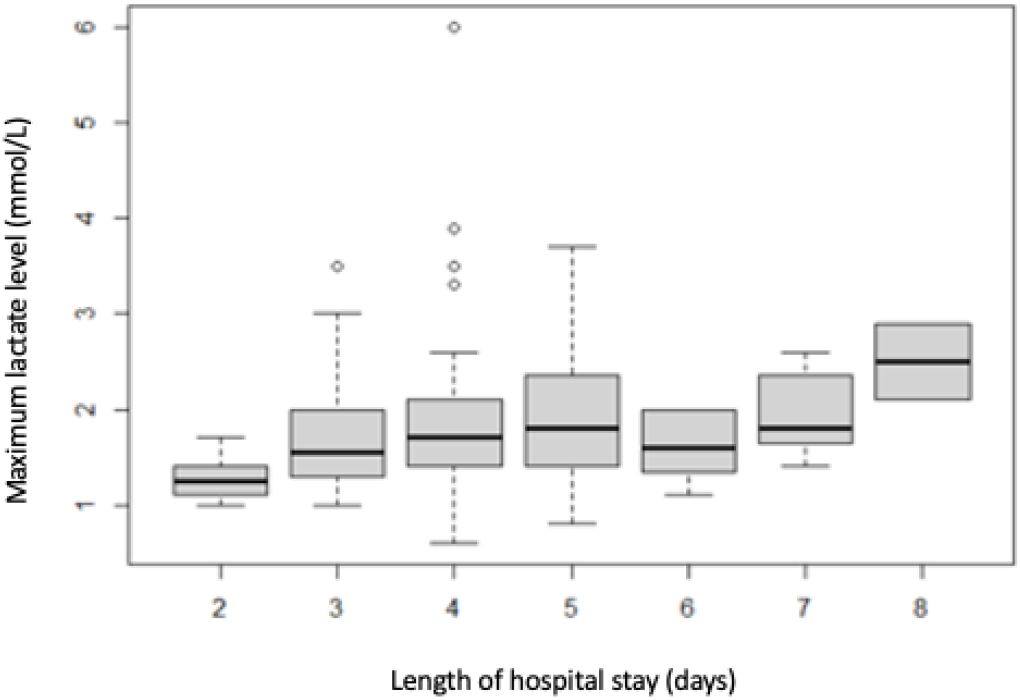
The box plot demonstrates the maximum lactate level (mmol/L) in study participants according to the hospital stay (days).

### Comparison between lactate and HCT improvement during fluid resuscitation

The reduction in lactate and corresponding HCT following fluid boluses (10ml/kg: maximum 500 ml) were evaluated to understand the response to fluid therapy. Fluid boluses received by study participants included crystalloid, dextran (colloid), both crystalloid/dextran combination or blood. There were 82 instances of receiving fluid boluses by study participants guided by the dengue management protocol [5]. Both venous lactate and venous HCT values were recorded immediately before and one hour after each fluid bolus. The lactate difference (pre-bolus lactate - post-bolus lactate) and the HCT difference (pre-bolus HCT - post-bolus HCT) for each instance of administering fluid bolus was plotted against the fluid type (Figure 3).

**Figure 3:**
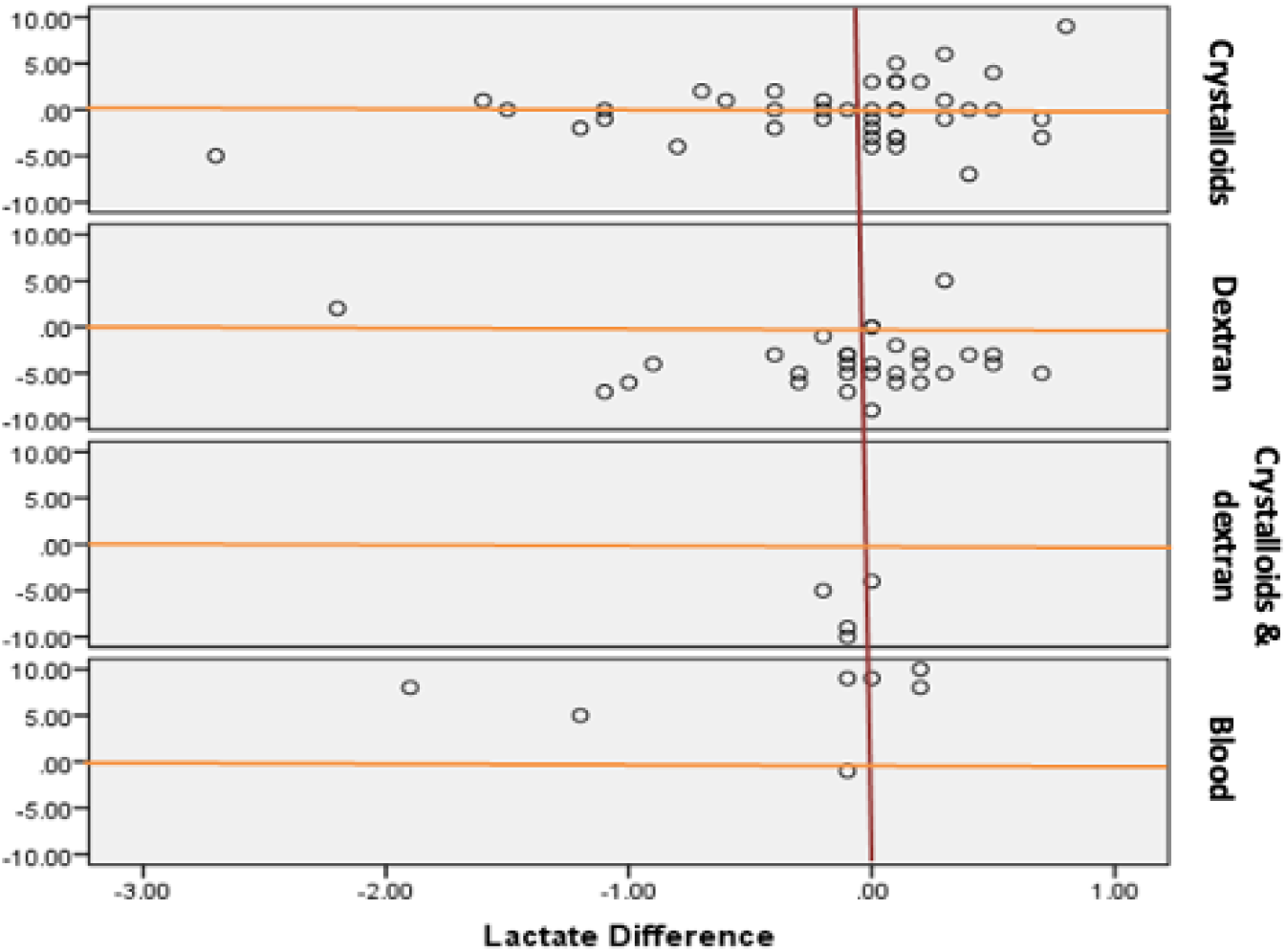
All instances of a study participant receiving a fluid bolus were evaluated to derive the lactate and HCT difference. The difference of lactate and HCT observed were plotted in the scatter plot. The X-axis represents the difference in the lactate and the Y-axis shows the difference in the HCT pre- and post-fluid bolus for each type of fluid. The red line represents value “0” with differences given as positive or negative values.

Results demonstrate that HCT reduction expected by administration of the fluid bolus was seen following dextran and crystalloid/dextran combination (Figure 3). This difference was statistically significant (Table 3). However, the expected reduction in HCT was not observed in most instances (16/22) following crystalloid bolus administration (Table 3). The HCT value increased in almost all instances of giving blood. In contrast, a statistically significant reduction in lactate was not observed following administration of any of the fluid boluses (Table 3).

**Table 3:** Comparison of the mean lactate and mean HCT difference after administration of fluid boluses.

| Type of fluid bolus administered | Mean lactate<br>difference | p-value | Mean HCT<br>difference | p-value |
| --- | --- | --- | --- | --- |
| Crystalloid | 0.185 | 0.096 | 0.073 | 0.880 |
| Blood | 0.414 | 0.225 | -6.857 | 0.003 |
| Dextran | 0.11 | 0.299 | 3.7 | 0.000 |
| Crystalloid/Dextran combination | 0.1 | 0.092 | 7 | 0.018 |

## Discussion

The usefulness of lactate as a bedside test to guide fluid therapy in DHF is under evaluated. This observational study was conducted to fill the gap in literature and to identify the correlation of venous lactate measured by a lactate meter and HCT during fluid resuscitation of DHF. Our study was conducted on a significant number of dengue patients in critical phase comparing the behavior of currently advocated HCT with lactate levels during fluid resuscitation. Majority of study participants were admitted to hospital on day three of the illness and majority entered critical phase on day four of the illness in keeping with the natural history of DHF. As expected, the study population was young with a mean age of 32 years and most of them did not have major comorbidities which could influence the outcome from dengue illness. Most prominent feature of fluid leakage was the presence of ascites followed by pleural effusion. The routine in-ward ultrasound scans have contributed to the early detection of effusions in the study setting. Only 24 (14.8%) patients showed evidence of a rise in HCT > 20% and this is an observation in previous studies as well due to patients receiving adequate fluids during the pre-critical phase of the illness [15].

In healthy individuals, lactate levels are less than 1.0 mmol/L when measured in both arterial and venous blood. Raised serum lactate level is known to correlate with higher mortality and morbidity in critically ill patients [16]. Moreover, it’s recommended to use lactate clearance during initial resuscitation in patients with sepsis [17]. In our study, the majority (n=154, 95.5%) of the study participants had a median lactate value between 0-2 mmol/L. Lactate levels > 2 mmol/L was shown to be associated with development of severe dengue and plasma venous lactate ≥2.5 mmol/L had a sensitivity of 90% and a specificity of 87.6 % in diagnosing severe dengue in previous studies [18]. A rise in lactate beyond 2 mmol/L was seen in only 8 (4.3%) study participants despite elevations of HCT beyond 45 seen in 43 (26.5%).

We analyzed 77 (out of 82) occasions where fluid boluses were given in the form of crystalloid, dextran, crystalloid/dextran combination or blood (Figure 3). The mean difference of lactate and HCT after a fluid bolus was calculated. We did not find a significant lactate difference after a fluid bolus. Meanwhile, there was a significant improvement of HCT after dextran and crystalloid/ dextran combination as expected after a bolus. Furthermore, an increase in HCT was seen after blood transfusion (Table 3). Interestingly, crystalloid boluses did not show a significant improvement of HCT depicting that HCT behavior is not reliable to see the adequacy of fluid resuscitation when crystalloids are used. The absence of a significant change in venous lactate following fluid boluses is likely due to the narrow range of lactate levels observed in our patients. Study participants had their venous lactate between 0-2 mmol/L and only eight patients had compensated shock (pulse pressure ≤ 20 mmHg) in the study. There were no patients with decompensated shock (systolic blood pressure <90mmHg or reduction of systolic blood pressure by 20% or mean blood pressure <60mmHg). This may explain the reason for the mean lactate difference not being significantly different following fluid resuscitation compared to HCT. Interestingly, the duration of hospital stay was longer when the lactate level was high. Our data suggest that venous lactate is not an appropriate marker to use to monitor response during fluid resuscitation in DHF unless patients have profound shock causing significant hyperlactataemia.

Liver involvement is common in dengue infections and acute liver injury leading to hepatic dysfunction is a well-known entity causing high mortality [19,20]. In our study, elevated liver enzymes signify that there is transaminitis during the leaking phase of DHF. Moreover, around 60% of the patients had ALT and AST levels higher than 3 times the upper limit of normal. There is emerging evidence to suggest that the disturbances of liver microcirculation resulting from the direct effects of the virus and other immunogenic mechanisms such as cytokine effect on the liver sinusoidal lining and the resulting inflammation could be contributing to the pathogenesis of severe dengue [21,22]. The effect of fluid resuscitation and fluid balance on liver dysfunction is not well studied. In our study, the correlation between individual peak values of AST, ALT and venous lactate was 0.16 and 0.07 respectively.

There were few limitations in the study. There was zero mortality in this patient population and only one patient received intensive care admission for a short period indicating less severe disease in the DHF population studied. Careful fluid management, relatively younger age of patients, absence of co-morbidities and mild nature of this epidemic could be the reasons for uncomplicated DHF observed in the study population. Although it was planned to recruit severe DHF patients for the rest of the study, we had to conclude the study prematurely due to the SARS-CoV-2 pandemic.

In conclusion, the present study does not demonstrate a significant correlation between venous lactate levels and HCT levels during the critical phase of DHF. Lactate levels are not significantly elevated in DHF patients unless profound shock and multi-organ failure is seen rendering venous lactate inappropriate for monitoring response to fluid resuscitation. Venous HCT is reliable to assess response to administration of colloid and blood but does not correlate with the expected outcome following crystalloid administration.

## Data Availability

All relevant data is included in the manuscript.

## Acknowledgements

We acknowledge the contribution of Dr Sakunika Neunehella Rodrigo and Dr Raihana Riyaz in data collection.

## Author contributions

DW and JW were involved in conceptualization, methodology, funding acquisition and writing the draft. DS and SV were involved in formal analysis. EM and AW were involved in providing resources and methodology. NP was involved in conceptualization, writing the original draft and reviewing. All authors have contributed to reviewing the draft manuscript.

## Data statement

All relevant data is included in the manuscript.

